# Viral load and timing of infection define neutralization diversity to SARS-CoV-2 infection

**DOI:** 10.1101/2022.06.15.22276432

**Authors:** Sho Miyamoto, Takeshi Arashiro, Akira Ueno, Takayuki Kanno, Shinji Saito, Harutaka Katano, Shun Iida, Akira Ainai, Seiya Ozono, Takuya Hemmi, Yuichiro Hirata, Saya Moriyama, Ryutaro Kotaki, Hitomi Kinoshita, Souichi Yamada, Masaharu Shinkai, Shuetsu Fukushi, Yoshimasa Takahashi, Tadaki Suzuki

## Abstract

Immunity to SARS-CoV-2 in COVID-19 cases has diversified due to complex combinations of exposure to vaccination and infection. Elucidating the drivers for upgrading neutralizing activity to SARS-CoV-2 in COVID-19 cases with pre-existing immunity will aid in understanding immunity to SARS-CoV-2 and improving COVID-19 booster vaccines with enhanced cross-protection against antigenically distinct variants. This study revealed that the magnitude and breadth of neutralization responses to SARS-CoV-2 infection in breakthrough infections are determined by upper respiratory viral load and vaccination-infection time interval, but not by the lineage of infecting viruses. Notably, the time interval, but not the viral load, may play a critical role in expanding the breadth of neutralization to SARS-CoV-2. This illustrates the importance of dosing interval optimization in addition to antigen design in the development of variant-proof booster vaccines.

**One-Sentence Summary:** Viral load and infection timing define the magnitude and breadth of SARS-CoV-2 neutralization after breakthrough infection.

## Main Text

The proportion of individuals in the population with antiviral immunity to severe acute respiratory syndrome coronavirus type 2 (SARS-CoV-2) has increased dramatically due to the surge in coronavirus disease 2019 (COVID-19) cases and extensive worldwide vaccine campaigns. Nevertheless, the cases continue to increase, causing significant morbidity and mortality worldwide, due to the emergence of variants of concern (VOCs) with varying levels of increased transmissibility and resistance to existing immunity; thus, highlighting the necessity to develop next-generation COVID-19 vaccines. These vaccines must induce a durable and broad breadth of protective immunity covering all SARS-CoV-2 variants (*1*). Many open questions remain about how to induce high quality immunity that suppresses viruses with distinct antigenicity. A better understanding of immune responses to SARS-CoV-2 infection will ultimately yield better vaccine designs.

The most recently emerged VOC, the Omicron (B.1.1.529 lineage) variant, has spread rapidly worldwide at an unprecedented pace and with rapidly expanding viral genome diversity (*2*). As of March 2022, the BA.1, BA.1.1, and BA.2 sub-lineages account for the major Omicron sub-lineages (*2*). BA.1 is characterized by approximately 30 amino acid mutations, three short deletions, and one insertion in the spike protein; 15 mutations are located in the receptor-binding domain (RBD) and induce evasion of humoral immunity induced by prior infection or vaccination (*3–5*). Although BA.1 and BA.1.1 are presently the predominant Omicron variants, the relative incidence of BA.2 has increased rapidly in many countries and regions (*6*). BA.2, containing 11 spike protein mutations different from BA.1, has dramatically altered viral immune evasion capabilities, raising a serious global public health concern as a potential source of viral surge. Despite the high capability for humoral immunity evasion seen in BA.1, cross-neutralizing activity against BA.1 is elicited in booster vaccinees (*3, 7–9*). Research groups, including our own, report that cross-neutralizing activity against BA.1 and BA.1.1 is elicited in COVID-19 vaccine breakthrough infections (*10–13*).

Notably, convalescent sera from patients with breakthrough infections show lower or more variable neutralizing antibody titers against BA.1 than in those receiving booster vaccinations. Moreover, the time interval from vaccination to infection determines the induction of cross-neutralization to BA.1 and BA.1.1 variants; a longer interval contributing a greater induction of cross-neutralizing antibodies (*10, 14*). In contrast to booster vaccination, where both the dose and interval between vaccinations are controlled, the time interval between vaccination and breakthrough infection is not controlled resulting in diverse humoral immune responses against SARS-CoV-2 variants in breakthrough infections. However, other factors, such as symptoms, the infected viral strain, and viral replication, are expected to impact the immune response to SARS-CoV-2 infection. Primary drivers of humoral immune responses to SARS-CoV-2 in breakthrough infections have not been fully elucidated. Research in this area will provide fundamental insight into better vaccine designs, especially for booster vaccines.

Herein, we evaluated relationships between the amount of cross-neutralization activity to SARS-CoV-2 variants at acute and convalescent phases, upper respiratory tract viral load, and vaccination-infection time interval in non-Omicron breakthrough infection, as well as other case characteristics, and identified the key drivers of the magnitude and breadth of humoral immune responses to SARS-CoV-2 variants in individuals with immune histories due to combinations of vaccination and breakthrough infection.

## Results and Discussion

### Upper respiratory viral load and serum cross-neutralizing activity

We performed virological characterization of 220 breakthrough SARS-CoV-2 infected individuals, diagnosed 14 days after their second vaccination, using upper respiratory specimens collected within four days after diagnosis or disease onset (interquartile range; 0-0 days) (Tables S1, S2). Spike-mutation detection polymerase chain reaction (PCR) and viral genome analysis showed that most infected viruses were Delta or Alpha variants (Table S1). Viral RNA loads and infectious viral titers in upper respiratory specimens were significantly higher in Delta infected individuals than in Alpha infected individuals (Fig. 1A). Viral RNA loads and infectious viral titers were positively correlated to the same extent in both variants (Fig. S1A), suggesting no difference in the relationship between infectious viral amount and viral RNA levels between the variants. These results suggest that Delta variant replicates more efficiently in the upper respiratory tract in breakthrough infected individuals, consistent with previous reports (*15, 16*).

**Fig. 1.**
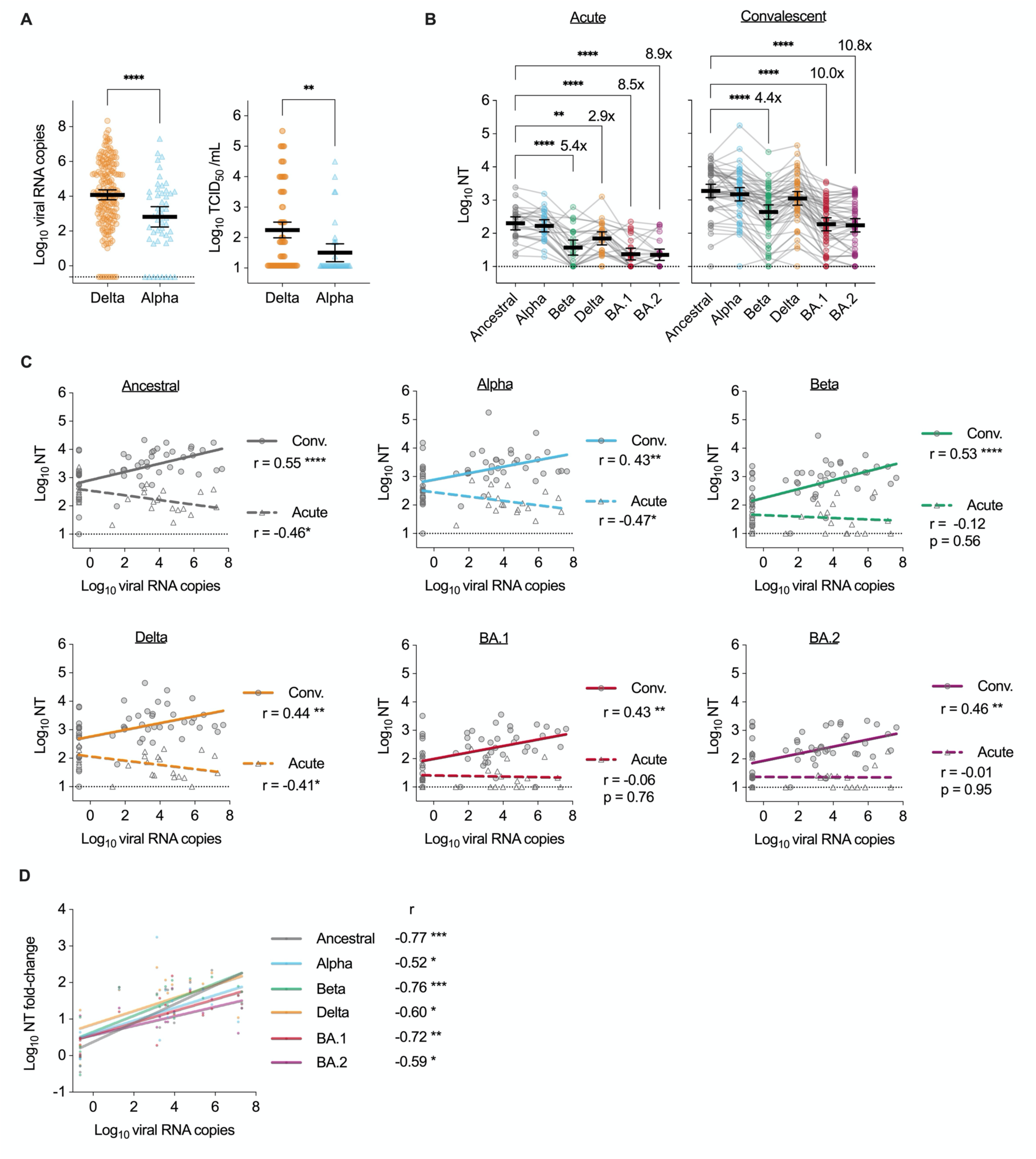
Relationship between humoral immune response and upper respiratory viral load in vaccine breakthrough SARS-CoV-2 infections. (**A**) Viral load in respiratory specimens at a near-diagnosis date. Viral RNA copies/reaction and viral titers for cases of SARS-CoV-2 Delta or Alpha variant infection were measured using reverse transcription quantitative polymerase chain reaction (RT-qPCR; left) and the median tissue culture infectious dose (TCID_50_; right), respectively. Viral titers are shown in viral isolation positive cases. Mean ± 95% confidence intervals (CIs) are presented. Viral loads were compared using unpaired t-tests. (**B**) Neutralization titers (NTs) in vaccine breakthrough case sera against SARS-CoV-2 pseudoviruses at acute and convalescent phases. Data from the same serum are connected with lines. Mean ± 95% CIs are presented for each serum titer. Titers between ancestral and newer variants were compared using ANOVA with Dunnett’s test. Fold-reductions are indicated above the columns. (**C**) Correlations between viral load in respiratory specimens at the near-diagnosis date and neutralization titers against the indicated variant at acute and convalescent phases. (**D**) Correlations between viral load and fold-changes in neutralization titers in acute and convalescent phases. (C, D) show regression lines, Pearson correlation r values, and significance levels. Statistical significance: \**p*<0.05, \*\**p* <0.01, \*\*\**p*<0.001, \*\*\*\**p*<0.0001. Dotted lines indicate cutoff values.

Sera from the enrolled cases were also obtained at one or two additional time points after infection. We defined the acute phase as within four days after diagnosis (four days after onset for cases diagnosed after onset) and the convalescent phase was defined as seven days after diagnosis or onset, based on the dynamics of anti-spike (S) RBD and anti-nucleoprotein (N) antibody titers (Fig. S1B). We excluded cases positive for anti-N antibody in the acute phase because they had a history of SARS-CoV-2 infection prior to breakthrough infection. Additionally, cases with acute phase anti-S RBD antibody titers <10 U/mL were excluded from this analysis because they were considered as low responders to COVID-19 vaccination. Following this, 26 cases with sera collected in the acute phase and 51 cases with sera collected in the convalescent phase (16 of whom had both acute and convalescent phase sera) were enrolled for further analysis. Serum neutralizing activity in these cases was measured using pseudovirus- or live virus-based assays (Figs. 1B, S1C). Neutralization titers to the ancestral and BA.2 strains were strongly correlated (Fig. S1D). Neutralization titers determined by pseudovirus-based assays were used for further analyses.

Neutralizing activity for all variants was higher in the convalescent phase than in the acute phase. (Figs. 1B, S1C). Acute phase sera showed lower neutralizing activity to Beta, Delta, BA.1, and BA.2 variants than to ancestral strain. Furthermore, convalescent phase sera showed lower neutralizing activity to Beta, BA.1, and BA.2 variants than to the ancestral strain. Notably, no obvious differences were found in neutralizing activity between BA.1 and BA.2 in both acute and convalescent phase sera, suggesting negligible antigenic difference between BA.1 and BA.2 in non-Omicron breakthrough sera. Acute and convalescent neutralization titers were lower to BA.2 than to ancestral strain (using live virus-based assays) (Fig. S1C), which confirmed the results obtained in the pseudoviruses-based assay. Convalescent phase sera from Delta breakthrough infections showed higher neutralizing activity against ancestral and Beta variants than sera from Alpha breakthrough infections, and both demonstrated equivalent neutralizing activity against Alpha and Delta variants. No apparent differences in neutralizing activity for BA.1 or BA.2 were observed between sera from Delta and Alpha breakthrough infections (Fig. S1E), suggesting that neutralizing antibodies against the infected variant in breakthrough cases were not necessarily induced preferentially over neutralizing antibodies against other variants.

To understand the impact of viral replication in breakthrough infections on antibody response, we analyzed the relationship between upper respiratory viral load and neutralizing activity to each variant in the acute or convalescent phase (Fig. 1C). In the acute phase, serum neutralization titers against ancestral strain and Alpha variant showed a significant negative correlation with upper respiratory viral load; similar trends were observed for acute phase serum neutralization titers against Delta variant; some serum neutralization titers against Beta, BA.1, and BA.2 variants in the acute phase were below detection limits, and precise correlations were not determined. However, convalescent phase serum neutralizing titers against all variants correlated positively with upper respiratory viral load. Notably, the relationship between upper respiratory viral load and acute phase neutralization was exactly opposite from the relationship between upper respiratory viral load and convalescent phase neutralization. Moreover, the increase in neutralizing activity against all variants from the acute to convalescent phases (using paired sera) was strongly positively correlated with upper respiratory viral load (Fig. 1D), indicating that lower levels of neutralizing antibodies during the acute phase of breakthrough infections are associated with higher viral replication levels in the upper respiratory tract, and that higher viral replication levels in the upper respiratory tract at the time of breakthrough infection induce a greater number of cross-neutralizing antibodies against a wide range of variants including Omicron variants in non-Omicron breakthrough infections. Further, it has been reported in non-human primate infectious models and observational studies on COVID-19 vaccine breakthrough infections that low levels of neutralizing antibodies prior to infection allow for efficient viral replication in the upper respiratory tract (*17–19*). This is consistent with the present results showing an obvious relationship between acute-phase neutralizing activity and viral load. It has also been reported that lower viral load in the upper respiratory tract tends to induce more neutralizing antibodies in unvaccinated SARS-CoV-2-infected patients (*20*), which is contrary to the present results showing a positive relationship between convalescent phase neutralizing activity and viral load in breakthrough infections. This suggests that the induction of neutralizing antibodies in breakthrough infections involves an interaction between preexisting immunity obtained through vaccination and viral replication during breakthrough infection.

### Cross-neutralizing activity

Previously, we reported that the cross-neutralizing activity of breakthrough convalescence sera against SARS-CoV-2 variants correlates with the vaccination-infection time interval (*10*). However, the induction of antiviral immunity is expected to involve additional factors, including age, sex, symptomology, the viral lineage, and viral replication. To explore the primary factors influencing neutralizing activity against each variant in breakthrough infections, we calculated correlation coefficients for each combination of factors: antibody titers, neutralization titers, age, sex, viral lineage (Delta, Alpha), viral load, the time interval from vaccination to infection, and the presence or absence of symptoms (Fig. 2A). In the convalescent phase, all neutralization titers were strongly and positively correlated. Most neutralization titers were also positively correlated in the acute phase. However, neutralization titers in acute phase sera did not correlate with neutralization titers in the convalescent phase sera for any variant (Fig. 2A), suggesting that neutralizing activity in the acute phase does not determine subsequent antibody responses in the convalescent phase. Additionally, sex, age, and symptom onset showed no clear correlation with neutralization titers in convalescent phase sera. Delta variant infection was positively correlated with neutralization titers to ancestral and Beta strains only, confirming the results shown above (Fig. S1E). Conversely, the vaccination-infection interval was positively correlated with neutralization titers to ancestral, Beta, BA.1, and BA.2 strains in convalescent phase sera. Additionally, upper respiratory viral load correlated strongly with neutralization titers against all variants in convalescent phase sera, suggesting that upper respiratory viral load at the time of diagnosis influences antibody responses in the convalescent phase following breakthrough infection.

**Fig. 2.**
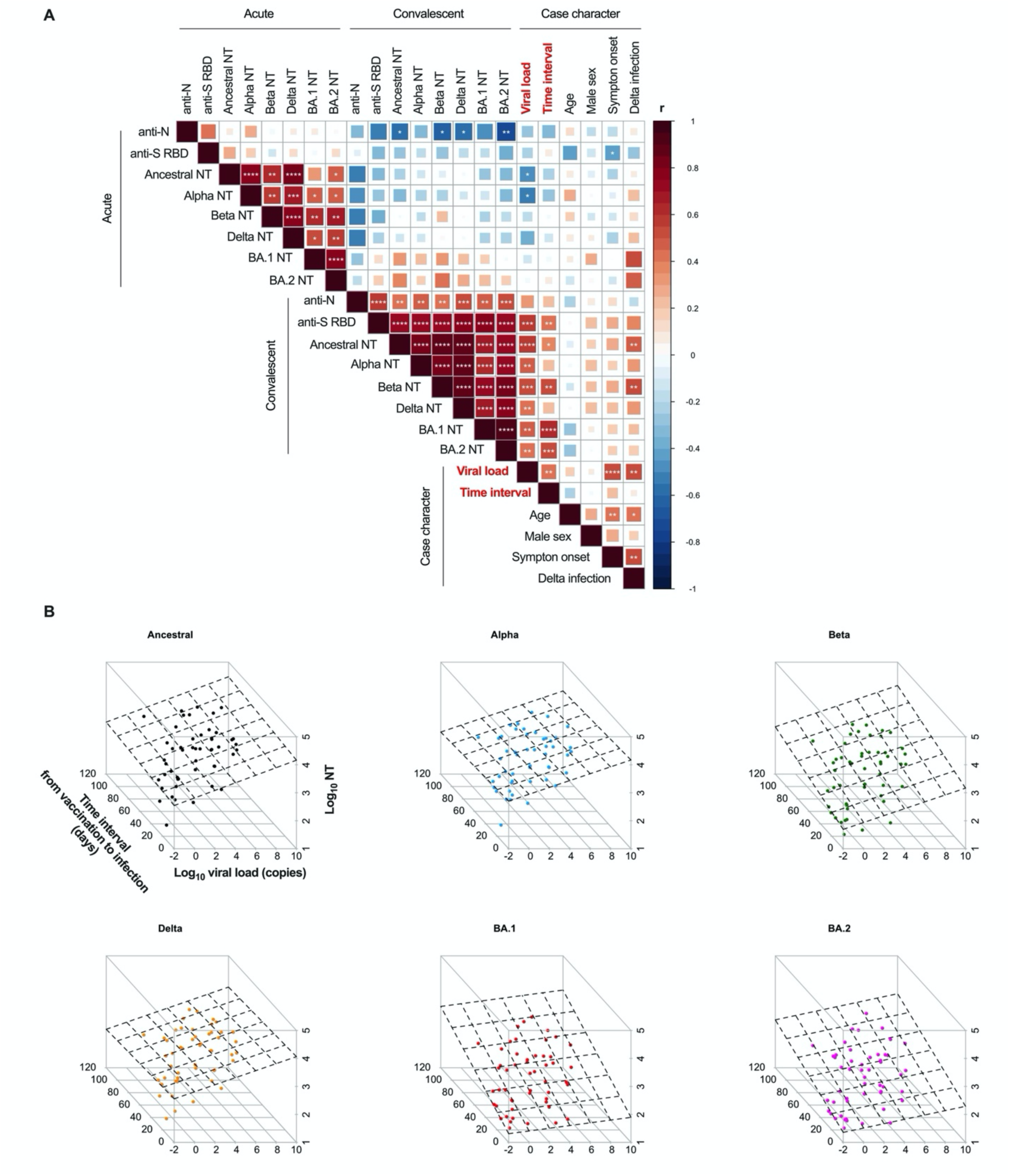
Determinants of neutralization serum titers against SARS-CoV-2 variants in breakthrough infections. (**A**) Spearman correlation matrix of antibody titers, viral load, and case characteristics for individuals with SARS-CoV-2 breakthrough infections. Neutralizing titers from pseudovirus neutralization tests, viral RNA copies used to represent viral load, and the time from vaccination to infection were employed herein. Male sex, disease onset, and Delta (vs. Alpha) variant infection were set as dummy variables (1 vs. 0). Spearman correlation r values were indicated using square size and a heat scale. The statistical significance level, corrected using the false-discovery rate (FDR), is shown in the square; * *p*<0.05, \*\**p*<0.01, \*\*\**p*<0.001, \*\*\*\**p*<0.0001. (**B**) Three-dimensional scatter plot of viral RNA load in respiratory specimens at a near-diagnosis date, the time interval from vaccination to infection, and neutralization titers against SARS-CoV-2 variants at the convalescent phase. The dotted planes represent regression planes. Table 1 shows the results of the multiple regression analysis relevant to the findings illustrated herein.

To independently evaluate the impacts of these two different factors on neutralizing activity, we performed a multiple regression analysis on three parameters: neutralization titers for each variant, viral load, and the vaccination-infection time interval (Table 1). Further we evaluated the slope-coefficients for viral load and vaccination-infection time interval on neutralization titers for each variant. The coefficients for viral load were significant for neutralization titers against ancestral, Alpha, Beta, Delta, and BA.2 strains (but not for BA.1). However, although the coefficients for vaccination-infection time interval were significant for neutralization titers against BA.1, and BA.2, they were not for the ancestral, Alpha, Beta, or Delta strains. Moreover, three-dimensional plots utilizing neutralization titers to each variant, viral load, and the vaccination-infection time interval as independent variables revealed that the regression plane skewed more towards viral load in neutralizing activity to the ancestral strain, Alpha, and Delta, and more towards the vaccination-infection time interval in neutralizing activity to Beta, BA.1, and BA.2 (Fig. 2B). These results suggest that upper respiratory viral load more strongly determines convalescent phase neutralizing activity to ancestral, Alpha, Beta, and Delta variants (vs. BA.1 and BA.2). Contrarily, the vaccination-infection time interval more strongly determined the convalescent phase neutralizing activity to BA.1, and BA.2. Two independent factors (upper respiratory viral load and the vaccination-infection time interval) showed differing magnitude of impact for each variant on the induction of variant neutralizing antibodies in breakthrough infections.

**Table 1.** Multiple regression analysis of neutralization titers, including viral load and the time interval from vaccination to infection.

| **Pseudovirus** | **Variable** | **Coefficient** | **95% CI** | **P-value** |
| --- | --- | --- | --- | --- |
| **Ancestral**<br>**(log<sub>10</sub> NT)** | Time interval (d) | 0.003 | 0.011, -0.006 | 0.542 |
|  | Viral load (log<sub>10</sub> copies) | 0.135 | 0.208, 0.062 | <0.001 |
| **Alpha** | Time interval | -0.002 | 0.008, -0.011 | 0.723 |
|  | Viral load | 0.122 | 0.203, 0.042 | 0.004 |
| **Beta** | Time interval | 0.008 | 0.018, -0.001 | 0.077 |
|  | Viral load | 0.123 | 0.202, 0.043 | 0.003 |
| **Delta** | Time interval | -0.002 | 0.008, -0.011 | 0.749 |
|  | Viral load | 0.126 | 0.206, 0.045 | 0.003 |
| **BA.1** | Time interval | 0.016 | 0.024, 0.008 | <0.001 |
|  | Viral load | 0.049 | 0.118, -0.021 | 0.166 |
| **BA.2** | Time interval | 0.011 | 0.02, 0.003 | 0.012 |
|  | Viral load | 0.078 | 0.152, 0.004 | 0.040 |

CI, confidence interval

### Antigenic distances

To better understand the overall picture regarding the potency of cross-neutralization, evaluating broadness of the neutralizing antigenicity is crucial. Antigenic differences among variants recognized by each serum were calculated as the distances between each variant and serum on antigenic maps (*21, 22*). Antigens and sera are positioned relative to each other on an antigenic map, and antigenic distances between antigens may vary depending on the nature of the sera used in the analysis. The distance between variants recognized as antigenically different is greater than that between variants recognized as antigenically similar. Additionally, a shorter distance from a serum to a variant on an antigenic map indicates higher neutralizing activity compared to other serum-variant pairs. Calculating the distance between each variant and serum on an antigenic map is useful for comparing the breadth of neutralizing activity for each serum. To evaluate how distances between variants and sera on an antigenic map change depending on nature of the sera, an antigenic map was generated using sera obtained from individuals who were vaccinated twice (2vax) or three times (3vax) with BNT162b2 (the Pfizer/BioNTech mRNA vaccine) (Fig. 3A).

**Fig. 3.**
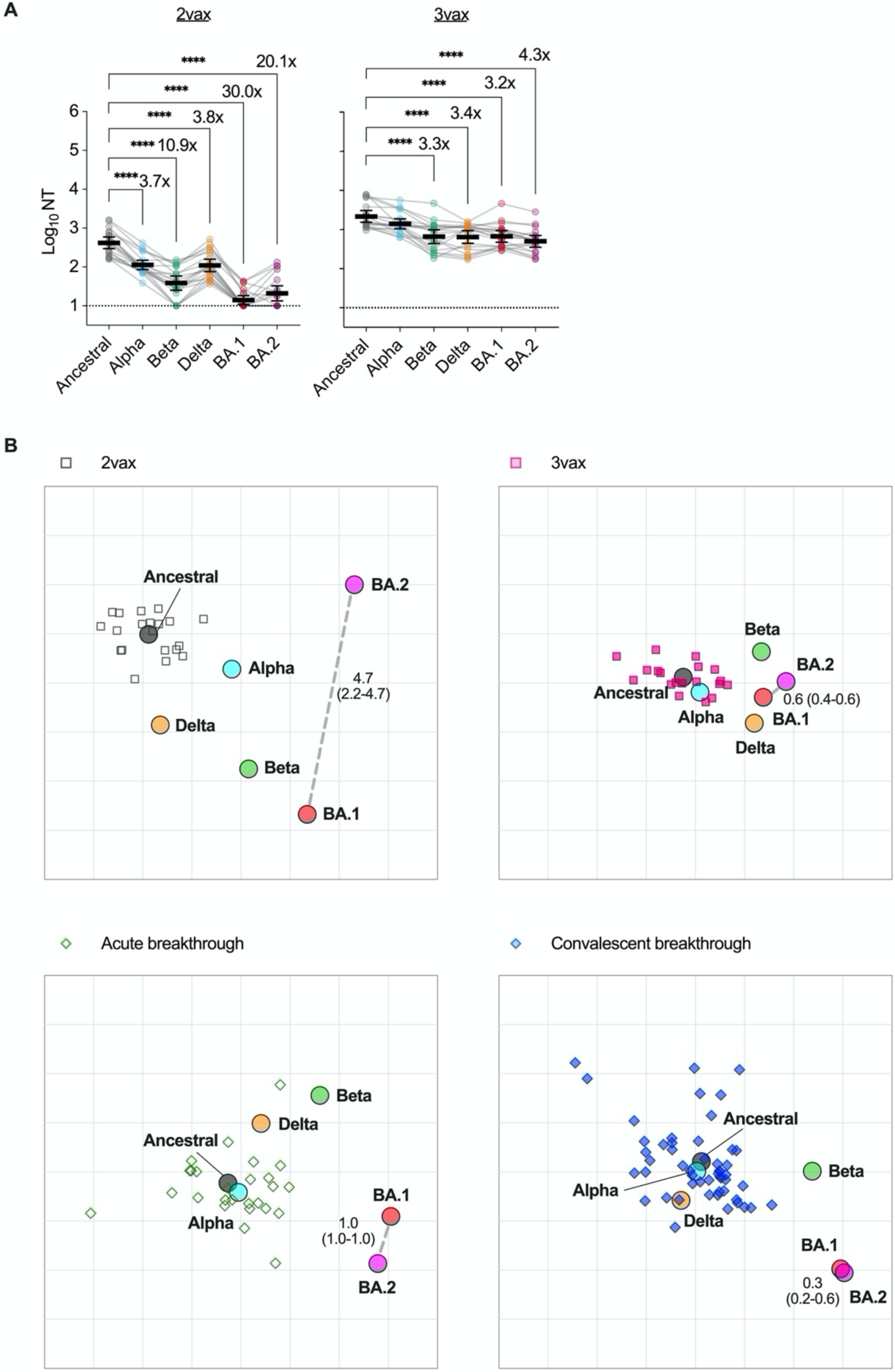
Antigenic maps of SARS-CoV-2 variants for each serum source. (**A**) Neutralization titers of sera obtained from individuals vaccinated twice (2vax) or three times (3vax) against variants of SARS-CoV-2 pseudoviruses. Data from the same serum are connected with lines, and the mean ± 95% CIs of each serum titer are presented. The titers between the ancestral and newer variants were compared using one-way ANOVA with Dunnett’s test; \*\*\*\**p*<0.0001. Fold reductions are indicated above columns given statistical significance. Dotted lines indicate cutoff values. (**B**) Antigenic cartography of each serum source for the acute and convalescent phases of breakthrough infection, 2vax, and 3vax (against SARS-CoV-2 variants). The variants are shown as circles and sera are indicated as squares or diamonds. Each square and diamond corresponds to the sera of one individual; colors represent the serum source. Each grid square (1 antigenic unit) corresponds to a two-fold dilution in the neutralization assay. Antigenic distance is interpretable in any direction. The median (interquartile rage) of the BA.1-BA.2 variant distance on the map is shown via gray dotted lines.

For 2vax sera, neutralization titers against Beta, BA.1, and BA.2 were more than 10-fold lower than titers against ancestral strain (Fig. 3A). The positions of each variant were scattered on the antigenic map, with variants being spaced far apart (Fig. 3B).

Conversely, for 3vax sera, there was only an approximately four-fold reduction in neutralization titers against Beta, BA.1, and BA.2 variants compared with that against ancestral strain (Fig. 3A). All variants (including BA.1 and BA.2) were spaced close together (Fig. 3B), suggesting that sera from individuals with booster dose vaccinations possesses an expanded breadth of neutralizing activity against SARS-CoV-2 variants (including BA.1 and BA.2), and confirming that distances between each variant on the antigen map vary in accordance with nature of the sera used in the analysis.

Antigen maps were generated using acute phase or convalescent phase sera from individuals with breakthrough infections (Fig. 3B). There were no obvious differences in the distances between variants in the acute and convalescent phases of breakthrough infection, and there was more variation in the serum positions in both the acute and convalescent phases in breakthrough infections than in maps depicting 2vax and 3vax sera, suggesting that the breadth of neutralizing activity in breakthrough sera is as diverse from case to case as the magnitude of neutralizing activity. Furthermore, the antigenic distances between the BA.1 and BA.2 variants on the antigenic maps generated from both the breakthrough infection and 3vax sera were shorter than those generated using 2vax sera (Fig. 3B), suggesting that a third exposure (due to vaccination or breakthrough infection) may have the effect of shrinking the antigenic differences between the variants recognized as antigenically distinct viruses by 2vax sera. The antigenicity of the BA.1 and BA.2 variants has been shown to differ in naïve infected hamster convalescence sera (*23*). Contrastingly, human observational studies have reported that sera from individuals receiving booster vaccines shows similar neutralizing activity against BA.1 and BA.2 variants, and that vaccine effectiveness in booster vaccinees did not differ between BA.1 and BA.2 variants (*24–26*). Moreover, a decrease in the antigenic distance between BA.1 and D614G variants in sera from booster vaccinees was reported in a prior study (*21*). These previous findings are consistent with our findings.

We noted that BA.2 spike introduced 11 amino acid mutations differing from BA.1 spike. The similarity in antigenicity between BA.1 and BA.2 variants recognized by 3vax and breakthrough infection sera suggests that the highly cross-neutralizing antibody induced by a third exposure (i.e., by vaccination or breakthrough infection) might overcome the effects of amino acid mutations specific to BA.2 and BA.1 spikes.

### Vaccination-infection time interval and breadth of neutralizing activity

To compare the neutralizing breadth for each serum (obtained from vaccinees with or without breakthrough infection), an antigenic map was generated using 2vax, 3vax, and breakthrough infection sera (Fig. 4A); 3vax sera tended to be located closer to Beta, BA.1, and BA.2 than 2vax sera (Fig. 4A). We calculated variant-serum distances between each serum to each variant. In the heatmap clustering individual variant-serum distances, Beta, BA.1, and BA.2 were clustered in an antigenic group (antigenic group 2; vs. antigenic group 1, which included ancestral strain, Alpha, and Delta variants) (Fig. 4B), suggesting that Beta, BA.1, and BA.2 are antigenically distinct from ancestral strain. Additionally, almost all 3vax sera (but not 2vax sera) were classified as type 3 serum, with a short variant-serum distance for all variants (Fig. 4B), suggesting that booster vaccination conferred a greater breadth of neutralizing activity against SARS-CoV-2 (including antigenically distinct variants). However, in breakthrough infection sera, neither acute nor convalescent phase sera were classified into a specific serum type (Fig. 4B), suggesting that the breadth of neutralizing activity in breakthrough infection was diversified from case to case.

**Fig. 4.**
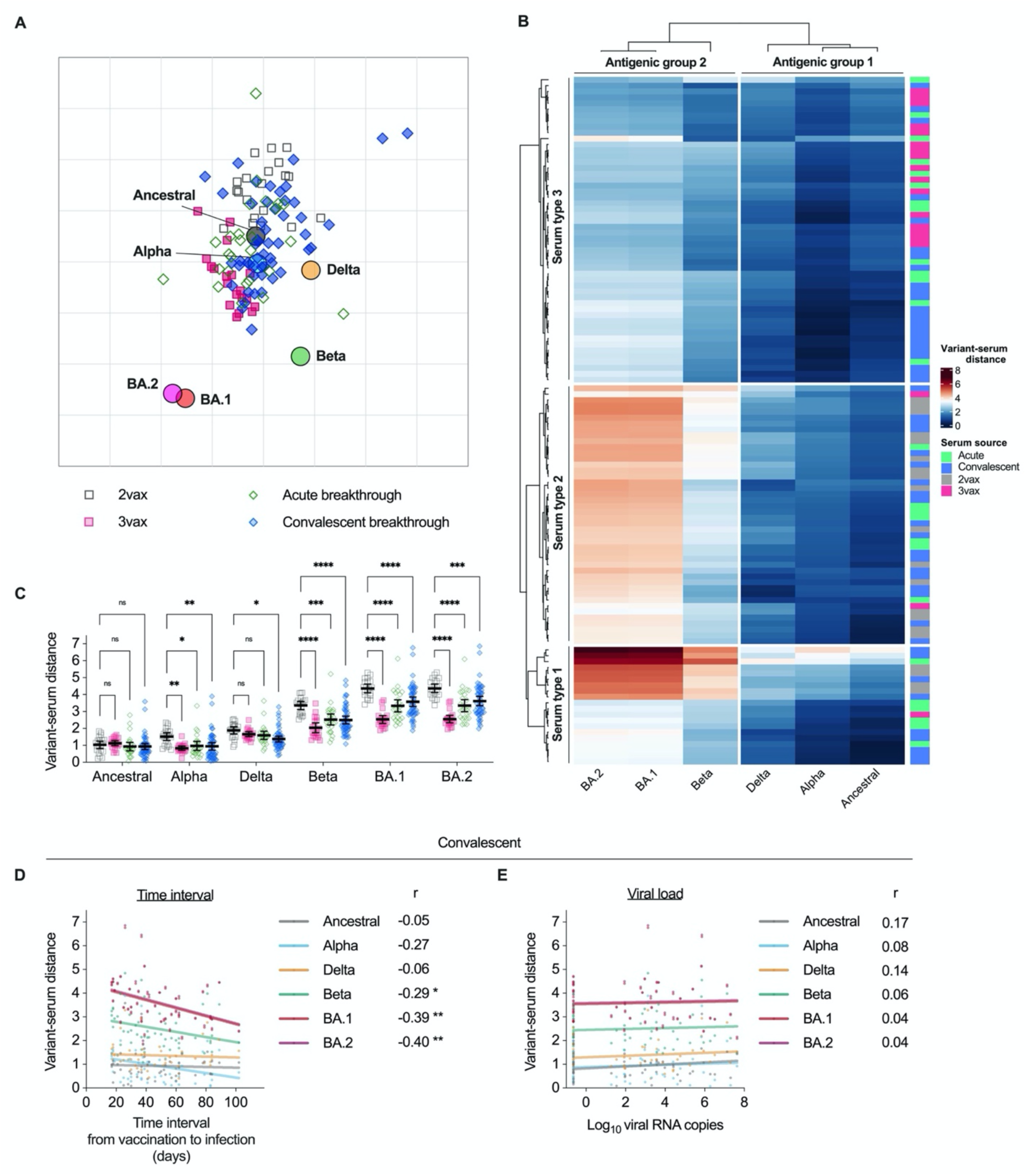
Characterization of heterogeneous serum populations based on antigenic map distance, and sources of variation. (**A**) Antigenic cartography of sera from acute/convalescent phases for breakthrough infections and for those receiving two (2vax) or three vaccines (3vax). Variants are shown as circles and sera are indicated as squares or diamonds. Each square and diamond corresponds to the sera of one individual; colors represent the serum source. Each grid square corresponds to a two-fold dilution in the neutralization assay. Antigenic distance is interpretable in any direction. (**B**) Clustering the antigenicity of SARS-CoV-2 variants and serum reactions against the variants. The heat map shows the variant-serum distance from the sera. Two antigenic groups and three serum types were divided by k-means clustering. The serum sources are indicated by a color bar on the right. (**C**) Comparison of variant-serum distance from the sera. Distances between the serum sources against 2vax sera were compared using two-way ANOVA with the Sidak test. (**D**) Correlation between the time interval from vaccination to infection and variant-serum distance at the convalescent phase. (**E**) Correlation between viral load in respiratory specimens at a near-diagnosis date and the variant-serum distance of sera at the convalescent phase. Statistical significance: ns, not significant, \**p*<0.05, \*\**p*<0.01, \*\*\**p*<0.001, \*\*\*\**p*<0.0001.

Next, variant-serum distances were compared for 2vax, 3vax, acute phase, and convalescent phase sera (Fig. 4C). A shorter variant-serum distance indicated a higher degree of neutralization. The variant-serum distances for the ancestral strain in 2vax, 3vax, acute phase, and convalescent phase sera were shorter than those for the other variants and did not differ among serum sources. Conversely, variant-serum distances for antigenically distinct variants, such as Beta, BA.1, and BA.2, were relatively longer than those for the ancestral, Alpha, and Delta variants. The variant-serum distances for 3vax, acute phase, and convalescent phase sera (in regard to the antigenically distinct variants) were statistically significantly shorter than for 2vax, suggesting that antibodies with an expanded breadth of neutralization covering antigenically distinct variants were induced in booster dose vaccinees in the acute and convalescent phases of breakthrough infection.

Following this, we evaluated the impact of the infecting viral lineage on variant-serum distances, and found no differences in variant-serum distance in either the acute or the convalescent phase (Fig. S2A). This suggests that the breadth of serum neutralization does not vary between Alpha and Delta breakthrough infections, and that Alpha and Delta breakthrough infections (antigenic group 1) equally impact the extent of the breadth of neutralization against different antigenic variants (antigenic group 2).

Moreover, to assess the effect of the time interval on the breadth of neutralization, breakthrough sera were divided into short (early breakthrough) and long (late breakthrough) time interval groups, and variant-serum distances were compared for each variant and serum. The variant-serum distances for BA.1 and BA.2 in the sera of the late breakthrough group were shorter than those in the early breakthrough group in convalescent phase sera (Fig. S2B). Intriguingly, the variant-serum distance did not change between the early and late breakthrough groups for either variant in acute phase sera (Fig. S2B). These observations suggest that the time interval from vaccination to infection strongly determines the breadth of neutralizing antibodies in the convalescent phase induced after breakthrough infection.

We evaluated the correlation between variant-serum distances in breakthrough infection sera, the vaccination-infection time interval, and the upper respiratory viral load (Figs. 4D, 4E). Variant-serum distances for the ancestral, Alpha, and Delta strains did not correlate with the vaccination-infection time interval, while the variant-serum distances for antigenically distinct variants correlated negatively. Upper respiratory viral load did not correlate with variant-serum distances for any variants. These results suggest that the vaccination-infection time interval acts as a primary determinant of variant-serum distances for antigenically distinct variants from the vaccine antigen and ancestral strain without being limited by the antigenicity of the infecting virus or viral replication, and that a longer time interval contributes to expanding the breadth of neutralization regarding antigenically distinct variants.

COVID-19-vaccine breakthrough infections were previously found to elicit robust cross-neutralizing antibody responses against several SARS-CoV-2 variants, which were largely recalled from memory B cells induced by previous vaccinations (*27*). mRNA vaccines induce a persistent germinal center B cell response in the draining lymph nodes at least 12 weeks after the second vaccine dose, enabling the generation of robust humoral immunity (*28*). Memory B cells recognizing Omicron and other variants are known to proliferate after the second vaccination (*29, 30*) and memory B cells recognizing the Omicron spike proliferate after the third vaccination (*31*). These findings support the contention that a third exposure increases the B cell population producing antibodies with high cross-neutralizing potency. Indeed, affinity maturation of IgG antibodies to the spike-protein-conserved region persisted for more than three months after SARS-CoV-2 infection in a prior study (*32*). Furthermore, cross-neutralization ability against antigenically distant variants was induced in a group with a longer first-to-second vaccination interval (*33*). Our study and previous findings support the importance of the time interval from vaccination in the progression of the breadth of humoral immunity to SARS-CoV-2 regarding both booster vaccinations and breakthrough infections.

Moreover, we showed that the upper respiratory viral load in Delta or Alpha (i.e., antigenic group 1) breakthrough infected individuals determined the resulting increase in the magnitude of the humoral immune response to SARS-CoV-2, including to variants belonging to antigenic group 2 (Fig. 1). This finding indicates that the viral replication level in the upper respiratory tract impacts neutralizing antibody production. Viral replication can modulate antiviral immunity through synthesizing viral antigens that serve as immunogens for inducing antiviral antibodies. Intriguingly, the effect of upper respiratory viral load on neutralizing antibody production in Delta/Alpha variant breakthrough infections was seen for both antigenically similar variants (ancestral, Alpha, and Delta variants) and antigenically distinct variants (Beta, BA.1, BA.2) across the antigenic barrier. This suggests that viral antigens supplied by upper respiratory viral replication in breakthrough infected cases might be sufficient to overcome the antigenic barrier by stimulating B cells against neutralizing epitopes that are conserved among variants in individuals infected after a sufficient time interval. Additionally, suggesting the potentially different approach to rational antigen design in booster vaccine development aimed at inducing a broad breadth of protective immunity (i.e., covering all SARS-CoV-2 variants). The time interval rather than the upper respiratory viral load was shown to expand the breadth of neutralization. However, further evaluation of the impact of upper respiratory viral load on the expansion of neutralization breadth in breakthrough infections with antigenically distinct viruses relative to the current vaccine strain is needed to fully elucidate the role of viral replication as a key driver of upgrading neutralizing activity in individuals with breakthrough infections.

Despite the overall strengths of this study, it has several limitations. First, the cases enrolled in this study were limited to breakthrough cases infected with the Alpha or Delta variant, and did not include cases infected with the Omicron variant. If the antigenicity of the Omicron variant is distinct from the current vaccine strain, we cannot rule out the possibility that viral load may affect neutralization breadth in breakthrough cases occurring due to Omicron variants. Second, the disease severity of the breakthrough cases enrolled in this study was predominantly biased toward asymptomatic and mild cases. However, COVID-19 disease severity is reported to be an important determinant of antibody induction in unvaccinated individuals (*20, 32*). Third, since acute phase sera from individuals with breakthrough infections showed a low neutralization titer (even against the ancestral virus) in our investigation, we could not determine an accurate fold reduction in the neutralizing antibody titer or an accurate correlation coefficient against respiratory viral load. Fourth, we did not assess T-cell immunity against SARS-CoV-2, which contributes to protection when antibody titers are low in non-human primate models (*19*) and may correlate with protection against severe disease. Fifth, there was no individual who experienced breakthrough infection after the booster dose enrolled this study. Sixth, breakthrough infection sera for individuals’ vaccination-infection time intervals of more than four months were not available in this study, and the interval that maximizes cross-neutralization potency remains unknown. Finally, our investigation did not evaluate the actual risk of reinfection by SARS-CoV-2 in individuals with a history of breakthrough infection, although there is evidence that neutralizing antibody titers can be a correlate of protection against ancestral strains and different variants (*34–36*).

### Concluding remarks

In conclusion, we found that non-Omicron breakthrough infections elicited robust cross-neutralizing activity against Omicron variants (including BA.2) across the antigenic barrier. Most importantly, our study clearly demonstrated that the length of the post-vaccination incubation period, but not the antigenicity of the infecting virus or the upper respiratory viral load, is a critical determinant in expanding the breadth of neutralization to antigenically distinct variants. Additionally, the upper respiratory virus load at the time of infection determines the amount of neutralizing antibody titer produced during the subsequent convalescent phases. Respiratory viral load and the vaccination-infection time interval each showed a variable magnitude of impact for each variant in regard to the induction of neutralizing antibodies in breakthrough-infected individuals. Consequently, antiviral immunity to SARS-CoV-2 is becoming increasingly diverse in individuals with breakthrough infections. Therefore, exploratory studies evaluating factors controlling the quality of the immune response after breakthrough infection can be a useful guidepost for the development of next-generation vaccines with the ability to induce a durable and broad breadth of protective immunity against all SARS-CoV-2 variants. This study highlights the opportunity to develop a variant-proof booster vaccine using an approach that does not rely on antigen design (*1*).

Moreover, it should be noted that control of upper respiratory viral replication in breakthrough cases is more difficult to achieve than control of the vaccination-infection time interval, as viral replication in the respiratory tract is influenced by multiple variables, including the infecting variant, amount of viral exposure, host susceptibility, treatment strategies (including antiviral drugs), innate immune response, and complex pre-existing acquired immunity to SARS-CoV-2. For this, inducing high quality antiviral immunity against SARS-CoV-2 following breakthrough infections is difficult to achieve. Therefore, breakthrough infection should not be considered as an option for boosting immunity.

## Data Availability

All data produced in the present study are available upon reasonable request to the authors

## Acknowledgments

We thank Akiko Sataka, Asato Kojima, Izumi Kobayashi, Yuki Iwamoto, Yuko Sato, Milagros Virhuez Mendoza, Noriko Nakajima, Kenta Takahashi at NIID for their technical support and Fukumi Nakamura-Uchiyama at Tokyo Metropolitan Bokutoh Hospital and Hidefumi Shimizu at JCHO Tokyo Shinjuku Medical Center for collection of blood samples from vaccinees. We also thank the following healthcare facilities, local health centers, and public health institutes for their contribution in providing us with valuable patient information and samples on breakthrough cases: Aki Health Center, Akiru Municipal Medical Center, Atsugi City Hospital, Akita Research Center for Public Health and Environment, Aomori Jikeikai Hospital, Aso Onsen Hospital, Chiba City Institute of Health and Environment, Chiba Prefectural Institute of Public Health, Chigasaki City Public Health Center, Chuhoku Branch Office for Public Health and Welfare, Daiwa Hospital (Osaka), Fukui Prefectural Institute of Public Health and Environmental Science, Fujimino Emergency Hospital, Fukuoka City Hospital, Gifu City Public Health Center, Gifu Prefectural General Medical Center, Gifu Prefectural Research Institute for Health and Environmental Sciences, Gunma Prefectural Institute of Public Health and Environmental Sciences, Gunma Saiseikai Maebashi Hospital, Hachioji City Public Health Center, Hakodate City Institute of Public Health, Hakodate Public Health Center, Harada Hospital (Hiroshima), Hyogo Prefectural Institute of Public Health Science, Ikegami General Hospital, IMS Fujimi General Hospital, IMS Sapporo Digestive Disease Center General Hospital, Inba Health and Welfare Center, International Goodwill Hospital, International University of Health and Welfare Hospital, International University of Health and Welfare Mita Hospital, Ishikawa Prefectural Central Hospital, Ishikawa Prefecture Health and Welfare Department, Ishikawa Prefectural Institute of Public Health and Environmental Science, Itami City Hospital, Japanese Red Cross Gifu Hospital, Japanese Red Cross Kanazawa Hospital, Japanese Red Cross Kumamoto Hospital, Japanese Red Cross Narita Hospital, JCHO Kanazawa Hospital, JCHO Nankai Medical Center, JCHO Takanawa Hospital, Juntendo University Hospital, Kaga Medical Center, Kameda Medical Center, Kasai Clinic (Osaka), Kashiwa Public Health Center, Kawachi General Hospital, Kawaguchi Seiwa Hospital, Keio University Hospital, Keiwakai Ebetsu Hospital, Kitakyushu Public Health Center, Kitakyushu Public Health Institute, Kitasato University Kitasato Institute Hospital, Kobe City Nishi-Kobe Medical Center, Kobe Ekisaikai Hospital, Komatsu Municipal Hospital, Koriyama City Public Health Center, Kumamoto City Hospital, Kumamoto City Public Health Center, Kumamoto Prefectural Institute of Public Health and Environmental Science, Kurume-shi Public Health Center, Kyoritsu Narashinodai Hospital, Kyoto City Institute of Health and Environmental Sciences, Kyoto Kujo Hospital, Kyoto Prefectural Institute of Public Health and Environment, Kyowakai Kyoritsu Hospital, Maebashi-shi Public Health Center, Makita General Hospital, Matsui Hospital (Tokyo), Matsumoto City Public Health Center, Mie Prefectunal Institute of Public Health and Environmental Sciences, Minami Kaga Health and Welfare Center, Minoh City Hospital, Misato Kenwa Hospital, Mitsui Memorial Hospital, Mizushima Kyodo Hospital, Nadogaya Hospital, Nagano City Public Health Center, Nagano City Public Health Institute, Nagano Environmental Conservation Research Institute, Nagasaki Prefecture Iki Hospital, Nagayama Hospital (Osaka), Nanbu Tokushukai Hospital, Nanshu Orthopedics Hospital, Narita Tomisato Tokushukai Hospital, National Center for Global Health and Medicine, National Hospital Organization Kyoto Medical Center, National Hospital Organization Osaka National Hospital, National Hospital Organization Nagoya Medical Center, Niigata City Health Center, Niigata Prefectural Institute of Public Health and Environmental Sciences, Nippon Medical School Chiba Hokusoh Hospital, Nishinomiya City Public Health Center, Nitta ENT clinic, Obihiro Daichi Hospital, Obihiro Health Center, Oita City Public Health Center, Oita Kouseiren Tsurumi Hospital, Oita Prefectural Institute of Health and Environment, Okayama Kyoritsu Hospital, Osaka Medical and Pharmaceutical University Hospital, Saiseikai Kanazawa Hospital, Saiseikai Moriyama Municipal Hospital, Saiseikai Yamaguchi Hospital, Saitama City Hospital, Saitama Nishi Kyodo Hospital, Sakura General Hospital (Aichi), Sakura Hospital (Kumamoto), Sapporo Public Health Office, Sasebo City General Hospital, Shibuya Clinic (Ishikawa), Shimane Prefectural Institute of Public Health and Environmental Science, Shimonoseki City Hospital, Shimonoseki Public Health Center, Shin Komonji Hospital, Shin-Yamanote Hospital, Shonan Daiichi Hospital, Suginami Public Health Center, Sumida Ward Public Health Center, Takasaki General Public Health Center, Tama Nambu Chiiki Hospital, Tamashima Central Hospital, Tochigi Prefectural Institute of Public Health and Environmental Science, Tokyo Medical and Dental University Medical Hospital, Tokyo Metropolitan Geriatric Hospital and Institute of Gerontology, Tokyo Metropolitan Institute of Public Health, Tokyo Women’s Medical University Adachi Medical Center, Tonan Hospital, Tsuchiya General Hospital, Tsukuba Central Hospital, Tsurukawa Sanatorium Hospital, Ueda Health and Welfare Office, Utsunomiya City Institute of Public Health and Environment, and Yamanashi Prefectural Institute for Public Health and Environment. We also thank GISAID for the platform to share and compare our data with data submitted globally.

## Funding

Japanese Society for the Promotion of Science Grants-in-Aid for Scientific Research (JSPS KAKENHI) grant 21K20768 (SMiyamoto)

Japan Agency for Medical Research and Development (AMED) grant JP21fk0108104 (TS), JP22fk0108637 (TS), JP20fk0108534 (YT), JP21fk0108615 (YT)

## Author contributions

Conceptualization: SMiyamoto, TA, TS

Methodology: SMiyamoto, TA, AU, TK, SS, HKatano, SI, SMoriyama, SF, YT, TS

Investigation: SMiyamoto, TA, AU, TK, SS, HKatano, SI, AA, SO, TH, YH, SMoriyama, RK, HKinoshita, SY, MS, SF

Data curation: SMiyamoto, TA, TS, SMoriyama, RK, MS

Formal analysis: SMiyamoto, TS

Visualization: SMiyamoto

Funding acquisition: SMiyamoto, YT, TS

Project administration: TS

Supervision: TS

Writing – original draft: SMiyamoto, TS

Writing – review & editing: SMiyamoto, TA, AU, TK, SS, HKatano, SI, AA, TH, YH, SMoriyama, RK, HKinoshita, SY, MS, SF, YT, TS

## Competing interests

Authors declare that they have no competing interests.

## Data and materials availability

All data are available in the main text or the supplementary materials. The SARS-CoV-2 viruses in this study are available from the National Institute of Infectious Diseases (NIID) under a material transfer agreement with the NIID (Tokyo, Japan).

## Materials and Methods

### Human participants and sampling

The characteristics of human participants included in this study are summarized in Table S1. Human plasma samples obtained from vaccinated health care workers with no infections who received two or three doses of BNT162b2 (Pfizer/BioNTech) mRNA vaccine were collected with written informed consent prior to enrollment and ethics approval by the medical research ethics committee of the National Institute of Infectious Diseases (NIID). Blood obtained from vaccinated health care workers was collected in Vacutainer cell preparation tubes (CPT; BD Biosciences, Franklin Lakes, NJ, US) and centrifuged at 1,800 x g for 20 min. Peripheral blood mononuclear cells (PBMCs) were suspended in plasma and harvested into conical tubes, followed by centrifugation at 300 x g for 15 min. The plasma was then transferred into another conical tube and centrifuged at 800 x g for 15 min, and the supernatant was transferred into another tube to completely remove PBMCs.

Human serum samples obtained from breakthrough cases were also included in this study. A breakthrough infection was defined according a positive result in a test detecting SARS-CoV-2 RNA or antigen conducted on a respiratory specimen collected ≥14 days after the second vaccine dose. Demographic information, vaccination status, and respiratory samples for determining the infecting variant in the breakthrough cases included in this report were collected as part of the public health activity led by NIID under the Infectious Diseases Control Law and were published on the NIID website in order to meet statutory requirements. Sera obtained from breakthrough cases were collected concurrently for clinical testing provided by the NIID (with patient consent), and neutralization assays for this report were performed using residual samples as a research activity with ethics approval by ethics approval from the medical research ethics committee of NIID and informed consent.

To examine neutralization, plasma and serum samples were heat-inactivated at 56°C for 30 min before use. The median dose interval between the first and second vaccine dose for both vaccinated uninfected individuals and breakthrough cases was 21 days (Table S1).

Respiratory specimens were collected through nasal swabs, nasopharyngeal swabs, or saliva samples. Quantitative reverse transcriptase polymerase chain reaction (RT-qPCR) assays were completed at NIID for all respiratory samples to confirm sample quality for genome sequencing and to quantify the viral RNA load.

### Ethical approval

All samples, protocols, and procedures described herein were approved by the Medical Research Ethics Committee of NIID for involving human participants (approval numbers 1178, 1275, 1316, and 1321).

### RNA extraction and RT-qPCR

RNA was extracted from respiratory samples using the Maxwell RSC miRNA Plasma and Serum kit (Promega, Madison, WI, USA). Quantification cycle (Cq) values (i.e., viral RNA loads) were measured by RT-qPCR using the QuantiTect Probe RT-PCR Kit (Qiagen, Hilden, Germany) targeting the SARS-CoV-2 nucleoprotein (N) region via an NIID-N2 primer/probe set (*37*). The thermal cycling conditions were as follows: 50°C for 30 min; 95°C for 15 min; and 45 cycles of 95°C for 15 s and 60°C for 1 min. The Cq values of samples judged to be negative were analyzed by substituting a Cq value of 45. Cq values were converted to viral nucleoprotein RNA copy numbers/reaction according to a previously reported simple regression line (*38*). Viral isolation and PCR screening for mutations of interest (N501Y, E484K, L452R) were attempted for NIID-N2 primer/probe set positive samples.

### SARS-CoV-2 viral RNA genome sequencing

For respiratory specimens with Cq values of less than 32, SARS-CoV-2 whole viral genome sequences were determined. A primer set modified from the ARTIC Network’s V1 primer set (https://github.com/ItokawaK/Alt_nCov2019_primers/tree/master/Primers/ver_N3) was used for conducting multiplex RT-PCR. A DNA library was constructed from the RT-PCR products using the QIAseq FX DNA Library Kit (Qiagen) as described previously (*39*), and was subjected to next-generation sequencing using the MiSeq System (Illumina, San Diego, CA, USA). Consensus sequences of the viral genome were obtained using the ARTIC field bioinformatics pipeline, following the ARTIC-nCoV-bioinformatics SOP-v1.1.0 (https://artic.network/ncov-2019/ncov2019-bioinformatics-sop.html, 2020). The consensus sequences were uploaded to the Global Initiative on Sharing All Influenza Data (GISAID; https://www.gisaid.org/).

### PCR screening for mutation detection

To support genome sequencing analysis to determine variants, PCR screening for N501Y, E484K, and L452R mutations was completed for all RT-qPCR positive samples. Two in-house PCR assays to detect N501Y and L452R mutations, respectively, had previously been developed at NIID for public health surveillance purposes. To screen for N501Y mutations in the spike (S) region, we designed primers [5’-CTTGTAATGGTGTTRAAGGTTTTAATTGT and 5’-GGTGCATGTAGAAGTTCAAAAGAAAG] and TaqMan MGB probes [for N501 detection: 5’-FAM-CCAACACCATTAGTGGGTTG-MGB; for 501Y detection: 5’-VIC-CCAACACCATAAGTGGGTTG-MGB]. To detect L452R mutations in the S domain, we likewise designed primers [5’-GCGTTATAGCTTGGAATTCTAACAATC and 5’-ATCTCTCTCAAAAGGTTTGAGATTAGAC] and TaqMan MGB probes [for L452 detection: 5’-FAM-ATTATAATTACCTGTATAGATTGT-MGB; for 452R detection: 5’-VIC-TTATAATTACCGGTATAGATTG-MGB]. The final concentrations of the primers and probes were 0.6 μM and 0.1 μM, respectively. The reagents and the reaction conditions for RT-qPCR were as described above. For the E484K mutation, a commercial primer/probe for E484K (Takara, Kusatsu, Japan) was used.

We defined SARS-CoV-2 variants as follows: (i) Alpha lineage, samples in which E484 and 501Y were detected; (ii) Delta lineage, samples in which the 452R was detected; and (iii) undetermined, samples in which the 484K was detected or mutation detections failed. There were no discordant results for these PCR assays in regard to viral genome sequencing in included cases until August 2021.

### Cells

HEK293T cells obtained from the American Type Culture Collection (Manassas, VA, USA) were cultured in Dulbecco’s modified Eagle medium (DMEM; Fujifilm, Tokyo, Japan) supplemented with 10% fetal bovine serum (FBS) at 37°C, and were supplied with 5% CO_2_. VeroE6/TMPRSS2 cells (JCRB1819, Japanese Collection of Research Bioresources Cell Bank; Osaka, Japan) were maintained in low glucose DMEM (Fujifilm) containing 10% heat-inactivated FBS (Biowest, Nuaillé, France), 1 mg/mL geneticin (Thermo Fisher Scientific), and 100 U/mL penicillin/streptomycin (Thermo Fisher Scientific) at 37°C; these cells were supplied with 5% CO_2_.

### Viral isolation and titration

Viral isolation was attempted for all RT-qPCR positive cases with available residual respiratory specimens, as described previously (*40*). Briefly, VeroE6/TMPRSS2 cells (*41*) (JCRB1819, JCRB Cell Bank) were seeded in 96-well flat-bottom plates, respiratory specimens mixed with DMEM supplemented with 2% FBS and Antibiotic-Antimycotic Solution (Thermo Fisher Scientific) were inoculated in duplicate, and the culture supernatant was changed to fresh medium one day post-infection (d.p.i.) and was incubated at 37°C and supplied with 5% CO_2_. On one and five d.p.i., a cytopathic effect was observed. After five days, the supernatant was collected and RT-qPCR using the SARS-CoV-2 direct detection RT-qPCR kit (Takara) was performed in order to confirm the propagation of SARS-CoV-2. The median tissue culture infectious dose (TCID_50_) in the residual specimens was determined for all viral isolation positive cases.

### SARS-CoV-2 virus

We used the SARS-CoV-2 ancestral strain WK-521 (lineage A, GISAID ID: EPI_ISL_408667) and the Omicron BA.2 variant TY40-158 (lineage BA.2.3, EPI_ISL_9595813) in this study; these variants were isolated using VeroE6/TMPRSS2 cells at NIID with ethics approval provided by the medical research ethics committee of NIID for studies conducted in human subjects (#1178). More specifically, to isolate viruses belonging to the Omicron BA.2 variant TY40-158 strain, respiratory specimens collected from individuals screened at airport quarantine stations in Japan and transferred to NIID for whole genome sequencing were subjected to viral isolation using VeroE6/TMPRSS2 cells at NIID.

### VSV pseudovirus production

The vesicular stomatitis virus (VSV) pseudovirus bearing SARS-CoV-2 spike protein was generated as previously described (*42*). Briefly, the spike genes of SARS-CoV-2 ancestral, Alpha, Beta, and Delta variants were obtained from viral RNA extracted from the SARS-CoV-2 WK-521 strain (ancestral strain), QHN002 (Alpha; lineage B.1.1.7, GISAID: EPI_ISL_804008), TY8-612 (Beta; lineage B.1.351, GISAID: EPI_ISL_1123289), and TY11-927 (Delta; lineage B.1.617.2, GISAID:EPI_ISL_2158617), respectively, by RT-PCR (PrimeScript II High Fidelity One Step RT-PCR Kit, Takara). The spike gene of the SARS-CoV-2 Omicron variant was obtained from RNA extracted from nasopharyngeal swab specimens from patients infected with the SARS-CoV-2 Omicron BA.2 variant (TY40-158) by RT-PCR, as described above. The cytoplasmic 19 aa-deleted SARS-CoV-2 spike genes were cloned into a pCAGGS/MCS expression vector. HEK293T cells transfected with expression plasmids encoding the SARS-CoV-2 spike genes for the ancestral, Alpha, Beta, Delta, BA.1, and BA.2 variants were infected with G-complemented VSVΔG/Luc. After 24 h, the culture supernatants containing VSV pseudoviruses were collected and stored at −80°C until further use.

### Electrochemiluminescence immunoassay (ECLIA)

Antibody titers for the ancestral spike (S) receptor binding domain (RBD) and nucleoprotein (N) were measured using Elecsys Anti-SARS-CoV-2 S (Roche, Basel, Switzerland) and Elecsys Anti-SARS-CoV-2 S (Roche) kits according to manufacturer instructions.

### VSV pseudovirus-based neutralization assay

Neutralization of SARS-CoV-2 pseudoviruses was performed as described previously (*10, 32*). Briefly, serially diluted serum (with five-fold serial dilution of serum from participants with second vaccinations, or eight-fold serial dilution of serum from breakthrough infected patients and participants with third vaccinations, starting at a 1:10 dilution) was mixed with an equal volume of VSV pseudovirus bearing SARS-CoV-2 spike protein and incubated at 37°C for 1 h. The mixture was then inoculated into VeroE6/TMPRSS2 cells seeded on 96-well solid white flat-bottom plates (Corning; Corning, NY, USA). At 24 h post-infection, the infectivity of the VSV pseudovirus was assessed by measuring luciferase activity using a Bright-Glo Luciferase Assay System (Promega) and a GloMax Navigator Microplate Luminometer (Promega). The reciprocal half-maximal inhibitory dilution (ID_50_) is presented as the serum neutralization titer.

### Live virus neutralization assay

Live virus neutralization assays were performed as described previously (*10, 32*). Briefly, serum samples were serially diluted (via two-fold dilutions starting from 1:5) in high-glucose DMEM supplemented with 2% FBS and 100 U/mL penicillin/streptomycin and were mixed with 100 TCID_50_ SARS-CoV-2 viruses (WK-521, ancestral strain; and TY40-158, Omicron BA.2 variant), followed by incubation at 37°C for 1 h. The virus-serum mixtures were placed on VeroE6/TMPRSS2 cells seeded in 96-well plates and cultured at 37°C with 5% CO_2_ for five days. After culturing, the cells were fixed with 20% formalin (Fujifilm) and stained with crystal violet solution (Sigma-Aldrich, St. Louis, MO, USA). Neutralization titers were defined as the geometric mean of the reciprocal of the highest sample dilution that protects at least 50% of the cells from a cytopathic effect from two to four multiplicate series. Since sera from individuals who suffered from breakthrough infections were limited in quantity, this assay was performed only once. All experiments using authentic viruses were performed in a biosafety level 3 laboratory at NIID.

### Antigenic cartography

Antigenic maps based on neutralization titers against SARS-CoV-2 pseudoviruses were created using the *Racmacs* R function (The R Project for Statistical Computing, Vienna, Austria) with 2,000 optimizations (*22, 43*). Each grid square (1 antigenic unit) corresponded to a two-fold dilution in the neutralization assay. Antigenic distance (i.e., map distance) was used as the variant-serum distance (https://acorg.github.io/Racmacs/articles/intro-to-antigenic-cartography.html). Medians and interquartile ranges of the distances between BA.1 and BA.2 variants were calculated by Pythagorean theorem using the coordinates of antigenic maps in optimization steps. Heatmaps were created to visualize variant-serum distances using the *ComplexHeatmap* R function.

### Statistical analysis

Data analysis and visualization were performed using GraphPad Prism 9.3.1 (San Diego, CA, USA) and R 4.1.2. Measurements below the detection limit, excluding Cq values, were converted to half the detection limit. For statistical analysis, unpaired t tests, one-way analysis of variance (ANOVA) with Dunnett’s test, and two-way ANOVA with the Sidak test were used to compare viral loads, antibody titers, and variant-serum distances. Pearson correlation coefficients were used to assess correlations between continuous variables. Statistical significance was set at *p*<0.05. In correlation matrix analyses, Spearman correlations between variables were calculated with false discovery rate (FDR) correction and were visualized using the *psych* and *corrplot* R functions, as described previously (*44*). Coefficients were indicated using square size and the aforementioned heat scale.

In multiple regression analysis, each neutralization antibody titer against ancestral and newer SARS-CoV-2 variants in convalescent sera was regressed against respiratory viral load (*x*_1_, viral RNA copies) at a near-diagnosis date as well as the time interval (*x*_2_, days) from vaccination to infection. The model for neutralization titer *y* for participant *i* can be written as follows:

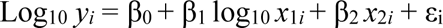

β_0_ represents the *y*-intercept; and β_1_, β_2_, and ε represent the slope coefficients and error term, respectively. These analyses and visualizations were performed using the *stats* and *scatterplot3d* R functions.

---

**Fig. S1.**
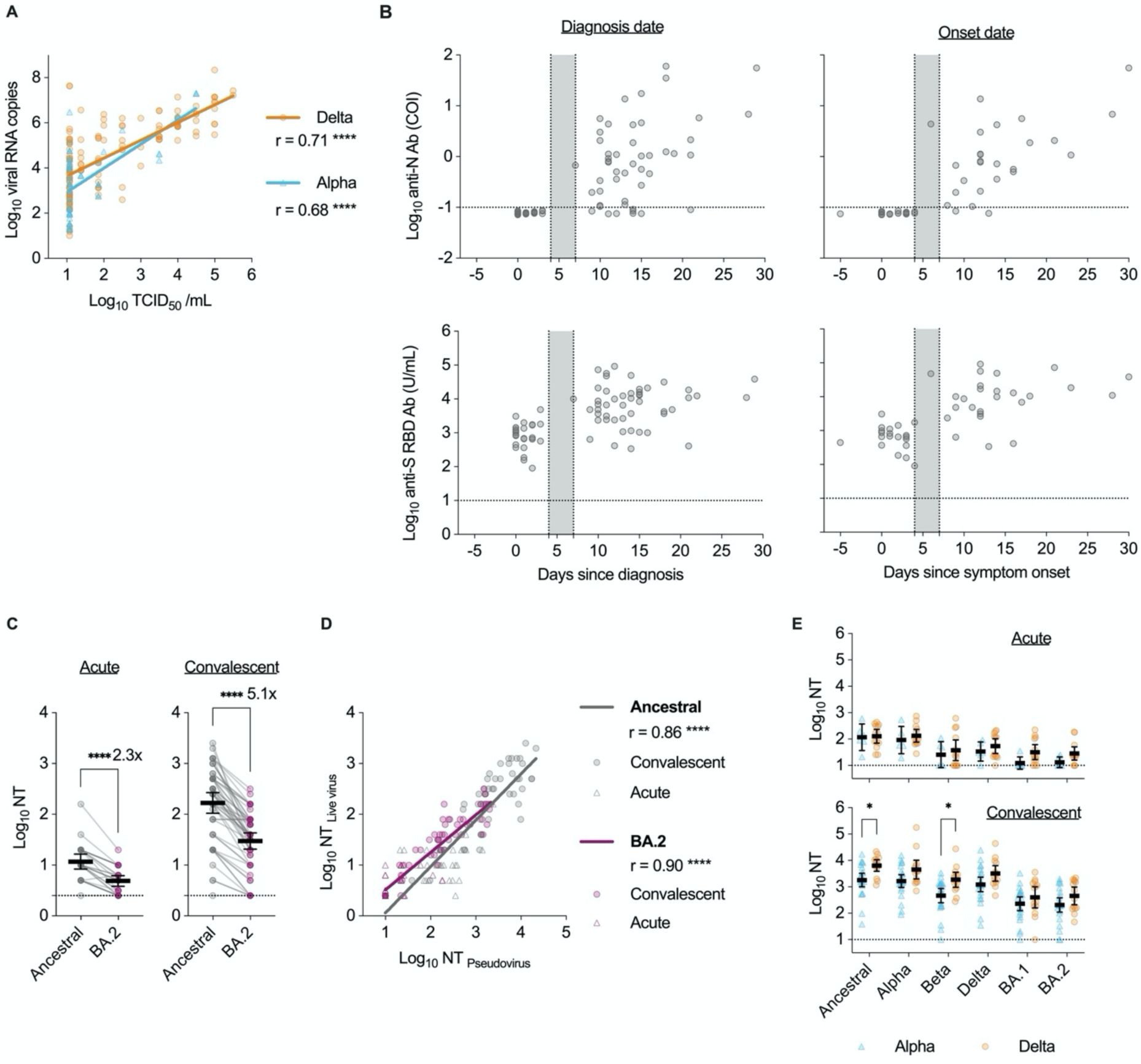
Supplemental characterizations of respiratory specimens and sera from individuals with SARS-CoV-2 vaccine-breakthrough infections. (**A**) Correlations between viral RNA copies and viral titers. (**B**) Anti-nucleoprotein (N) and anti-spike receptor binding domain (S RBD) antibody titers in breakthrough cases by days from diagnosis and days from symptom onset. Gray areas indicate 4-7 days since diagnosis or onset. (**C**) Neutralization titers (NTs) of breakthrough case sera against variants of SARS-CoV-2 live viruses at acute and convalescent phases. Data from the same serum are connected with lines, and means±95% confidence intervals (CIs) for each serum titer are presented herein. Titers were compared between ancestral and BA.2 variants using unpaired t-tests. Fold reductions are indicated above the columns. (**D**) Correlations of neutralization titers against pseudoviruses and live viruses. (**E**) Comparisons of neutralization titers between background variant types in breakthrough infections against SARS-CoV-2 pseudovirus variants. Titers were compared between background variant types using two-way analysis of variance (ANOVA) with the Sidak test. (A, D) Regression lines, Pearson correlation *r* values, and statistical significance levels are shown herein. Statistical significance: \**p*<0.05, \*\**p*<0.01, \*\*\**p* <0.001, \*\*\*\**p*<0.0001. Dotted lines indicate cutoff values.

**Fig. S2.**
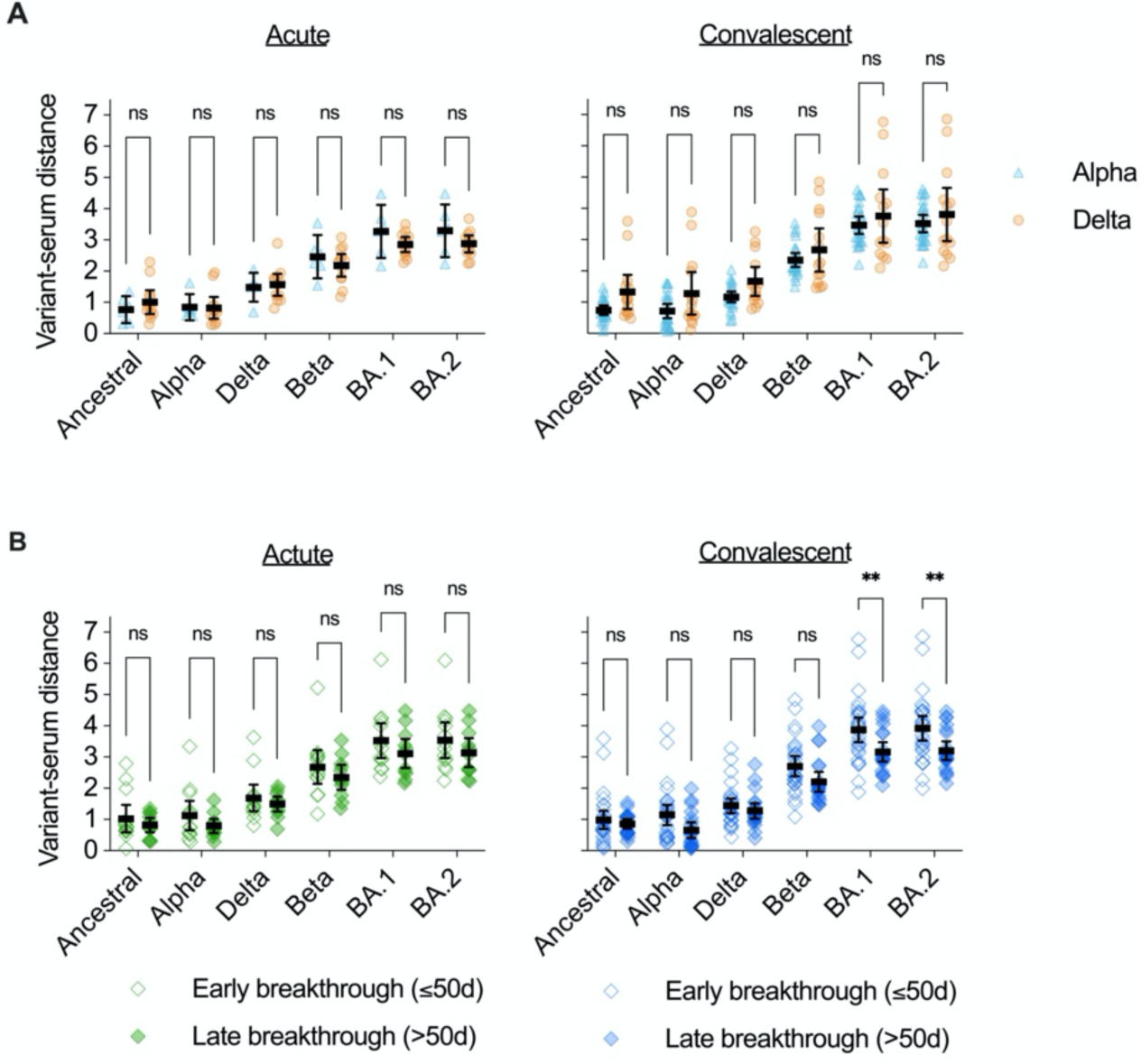
Effects of infection with SARS-CoV-2 variants and the timing of breakthrough infections on variant-serum distance. Variant-serum distances at acute and convalescent phases were calculated from the antigenic map shown in Fig. 4A. (**A**) Comparisons of variant-serum distances between background variant types in breakthrough infections at acute and convalescent phases. (**B**) Comparisons of variant-serum distances according to the timing of breakthrough infections at acute and convalescent phases. Early and late breakthrough infections were defined based on a demarcation at 50 days after vaccination. Distances were compared using two-way analysis of variance (ANOVA) with the Sidak test. Statistical significance: ns, not significant, \**p*<0.05, \*\**p*<0.01, \*\*\**p*<0.001, \*\*\*\**p*<0.0001.

**Table S1.** Medical and demographic characteristics of the participants included in this study.

| Sample | Breakthrough infection |  |  | 2 vaccination | 3 vaccination |
| --- | --- | --- | --- | --- | --- |
|  | Serum<br>(acute) | Serum<br>(convalescent) | Respiratory<br>specimen | Plasma | Plasma |
| N | 26 | 51 | 220 | 20 | 19 |
| Age (y) | 58 (35, 78) | 36 (26, 58) | 50 (30, 74) | 42 (32, 48) | 34 (31, 40) |
| Male sex | 17 (65%) | 35 (69%) | 141 (64%) | 4 (20%) | 9 (47%) |
| **Vaccine (2 doses)** |  |  |  |  |  |
| BNT162b2 | 24 (92%) | 50 (98%) | 189 (86%) | 20 (100%) | 20 (100%) |
| Not listed | 2 (8%) | 1 (2%) | 31 (14%) | 0 (0%) | 0 (0%) |
| **Severity** |  |  |  |  |  |
| Asymptomatic | 6 (21%) | 26 (46%) | 64 (28%) | NA | NA |
| Mild | 19 (68%) | 29 (51%) | 143 (63%) | NA | NA |
| Moderate* | 3 (11%) | 2 (4%) | 21 (9%) | NA | NA |
| Dose 1 to dose 2 interval (days) | 21 (21, 21) | 21 (21, 21) | 21 (21, 21) | 21 (21, 21) | 21 (21, 21) |
| Infection to sera collection (days) | 1 (0, 2) | 14 (11, 16) | NA | NA | NA |
| Infection to respiratory specimen collection (days) | 0 (0, 2) | 0 (0, 0) | 0 (0, 0) | NA | NA |
| Dose 2 to infection (days) | 46 (29, 62) | 41 (30, 62) | 50 (31, 73) | NA | NA |
| Dose 2/3 to sera collection (days) | 48 (32, 62) | 64 (44, 75) | NA | 32 (29, 34) | 35 (35, 35) |
| Dose 2 to dose 3 interval (days) | NA | NA | NA | NA | 245 (245, 246) |
| **Infection lineage** |  |  |  |  |  |
| Alpha | 6 (23%) | 24 (47%) | 50 (23%) | NA | NA |
| Delta | 12 (46%) | 14 (27%) | 136 (62%) | NA | NA |
| R.1 | 0 (0%) | 0 (0%) | 2 (1%) | NA | NA |
| Undetermined | 8 (31%) | 13 (25%) | 32 (15%) | NA | NA |
| Period of infection | Apr. 10 -<br>Aug. 20, 2021 | May 14 -<br>Aug. 20, 2021 | Apr. 10 -<br>Aug. 25, 2021 | NA | NA |

Median (interquartile range); n (%)
\*Moderate includes individuals who required oxygen

**Table S2.**

Individual characteristics, upper respiratory viral loads, and serum titers of the participants included in this study.

## Notes

### Competing Interest Statement

The authors have declared no competing interest.

### Funding Statement

This study was funded by Japanese Society for the Promotion of Science Grants-in-Aid for Scientific Research (JSPS KAKENHI) grant 21K20768 (SMiyamoto), and Japan Agency for Medical Research and Development (AMED) grant JP21fk0108104 (TS), JP22fk0108637 (TS), JP20fk0108534 (YT), JP21fk0108615 (YT).

### Author Declarations

The Medical Research Ethics Committee of National Institute of Infectious Diseases gave ethical approval for this work

